# The human cortical motor map in post-stroke hemiparesis in the presence of treatment-related clinically important recovery

**DOI:** 10.1101/2024.07.01.24309575

**Authors:** Tomoko Kitago, Joshua Silverstein, Michael B. Gerber, Charlene Thomas, Linda M. Gerber, Gary W. Thickbroom, Dylan J. Edwards

## Abstract

**Objectives:** To determine if intervention-related clinical improvement in post-stroke hemiparesis is associated with enlarged primary motor cortex representation.

**Methods:** Data were analyzed from a single site subset of the NICHE trial. Transcranial magnetic stimulation (TMS) motor mapping was performed in 23 participants (3-12 months post-stroke, 10 female, 40-88 yrs, baseline Fugl-Meyer Upper Extremity (FM-UE) range 13-58) before and after intervention. TMS motor maps were acquired for the extensor digitorum communis muscle (EDC) bilaterally, at 110% resting motor threshold (RMT).

**Results:** Improvement on the primary outcome measure (FM-UE) was statistically and clinically significant (mean pre= 38(±15), post= 45(±16); p <0.001; n=23). Significant improvement was also observed on secondary impairment and activity outcome measures (p<0.05). Ipsilesional hemisphere RMT and map volume (MV) remained unchanged (RMT pre = 42(±13), post = 41(±11), p=0.60; MV pre =77.98(±71.37) mV*mm^2^, post =109.54(±139.06) mV*mm^2^, p=0.36). The magnitude of clinical benefit was unrelated to difference in map characteristics from pre to post (Spearman’s rho= 0.06, p=0.79).

**Conclusions:** Intervention-related clinical improvement of the upper limb 3-12 months following stroke, was not associated with change in motor cortex excitability or increase in motor maps. Clinical improvements may not solely rely on consistent changes in the cortical motor representation.

## INTRODUCTION

Cortical reorganization has been proposed as a mechanism underlying recovery of upper limb motor deficits after stroke^1^. Changes in the area that is activated by transcranial magnetic stimulation (TMS, motor map) and the summed amplitude of the motor-evoked potentials (MEPs) over the region (map volume) are considered to reflect reorganization of the cortical areas controlling a specific muscle. Several studies have demonstrated expansion of TMS motor maps in association with clinical improvements in patients undergoing rehabilitative therapy after stroke^2,3^, suggesting that expansion of the cortical representation plays a role in therapy-induced improvements.

We acquired TMS motor maps longitudinally in a subset of participants enrolled in the multi-center Navigated Inhibitory rTMS to Contralesional Hemisphere (NICHE) randomized, controlled trial^6^. Participants underwent upper extremity task-oriented training over a six-week period (18 sessions), with each session preceded by 1 Hz active or sham repetitive TMS (rTMS). In the overall study, both active and sham rTMS groups improved significantly on upper extremity motor outcomes.

We hypothesized that clinical motor improvement would be accompanied by an increase in motor representation as measured by TMS motor maps in the ipsilesional hemisphere. We further predicted a positive correlation between TMS motor maps and changes in clinical status after training.

## METHODS

### Participants

A subset of participants from the NICHE study^6^ who underwent clinical assessments and TMS motor mapping before and after the six-week study intervention were included in this analysis. Participants had a unilateral ischemic or hemorrhagic stroke 3 to 12 months prior to enrollment with residual upper extremity weakness. Participants were excluded from analysis if mapping data was not available at either timepoint, due to a missed visit or inability to obtain a motor map. All participants gave written informed consent for the study, which was approved by the Institutional Review Board of the Burke Rehabilitation Hospital.

### Clinical Outcomes

The primary clinical outcome was change in motor impairment (Fugl-Meyer Upper Extremity Motor Assessment, FM-UE^7^; responders defined with >5-point improvement, considered the minimal clinically important difference)^8^. Secondary measures included upper limb activity limitation assessments (Action Research Arm Test, ARAT^9^, and Wolf Motor Function Test, WMFT)^10^.

### Transcranial Magnetic Stimulation

TMS mapping (110% RMT: extensor digitorum communis, EDC: left and right hemisphere), was conducted with Nexstim™ neuronavigation system. A target was placed on the location of the estimated hotspot, and a targeting grid (3 × 3 mm) was overlaid over the surface of the cortex. At least one pulse was delivered in each square of the grid that was mapped, and an effort was made to elicit at least two valid resting MEPs per square. Full details of the TMS procedures are described elsewhere^6^ and in the Supplement.

#### Mapping Analysis

MEPs were individually inspected and analyzed offline. Artifacts, active MEPs (defined as EMG activity >25μv within 100ms prior to the TMS pulse), and MEPs with a latency >25ms or <7ms were excluded from analysis. Motor mapping data were quantified using *NeuroMeasure*, an open source interactive software program^11^. Single-point mapping data was interpolated into a continuous function using a piecewise cubic spline curve fitting algorithm. Map area and map volume (spatial extent weighted by MEP amplitude, μV*mm^2^), was calculated for Pre and Post motor maps in each hemisphere^11^.

#### Statistical Analysis

Statistical analyses were performed in R Version 4.2.2. Wilcoxon signed-rank test was used to compare differences in outcome measures between time points. The relationship between the changes in map volume and the changes in clinical outcomes was examined using Spearman’s rank correlation. All p-values are two-sided with statistical significance evaluated at the 0.05 alpha level. As this study was exploratory, p-values were not corrected for multiple comparisons.

## RESULTS

Demographics and clinical characteristics of the 23 participants are reported in Table 1. There was no difference in outcomes between participants who received active (n=15) and sham (n=8) rTMS groups, thus they were combined for analysis in the present study. There were significant improvements in FM-UE from Pre to Post, with a mean increase in FM-UE score of 7.1 (SD 4.8) points, *p* < 0.001 (Figure 1A, Table 1). Secondary outcomes of upper limb activity (ARAT, WMFT) also improved significantly (Table 1).

**Table 1.**
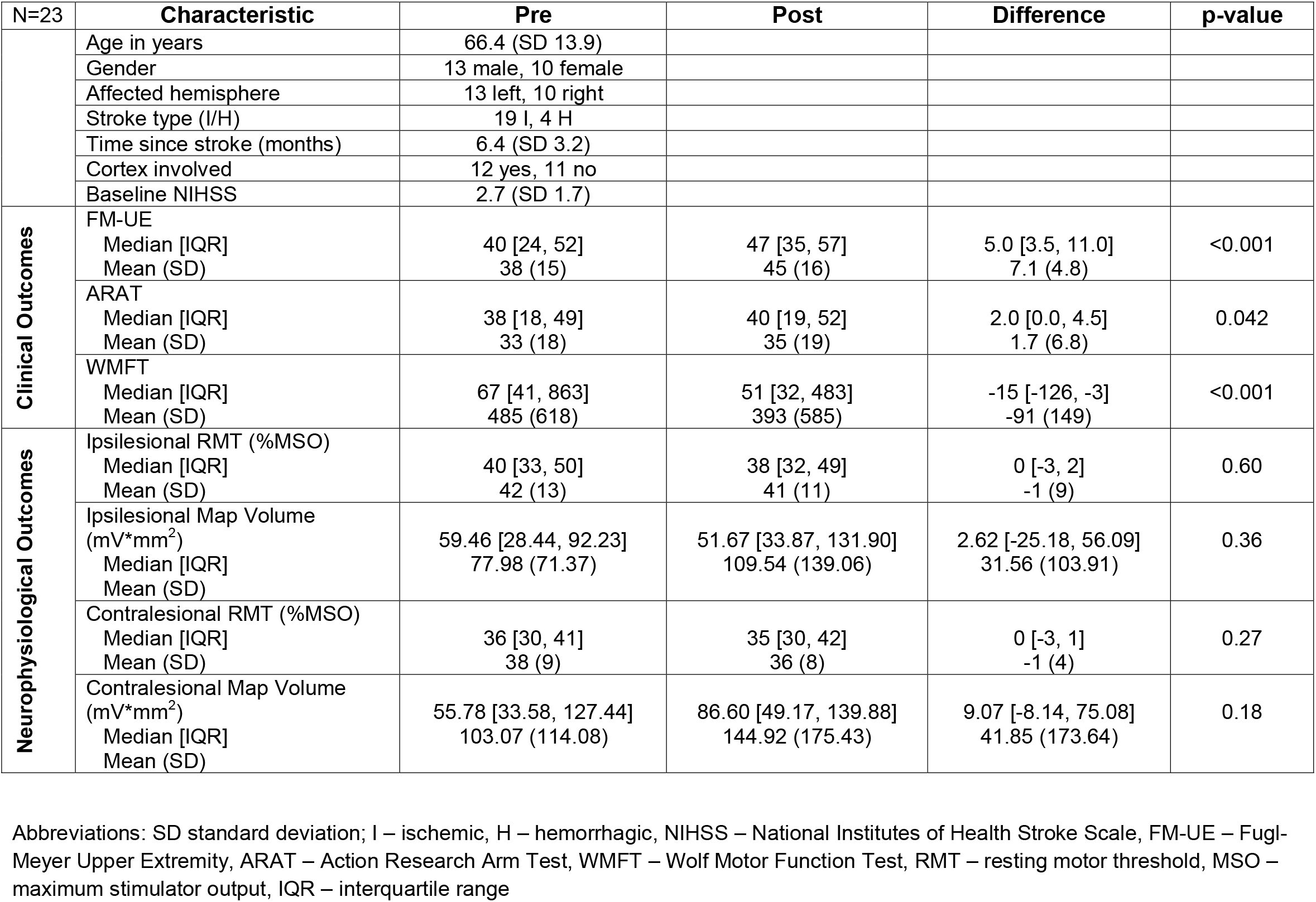
Study population and clinical and neurophysiological outcomes.

**Figure 1.**
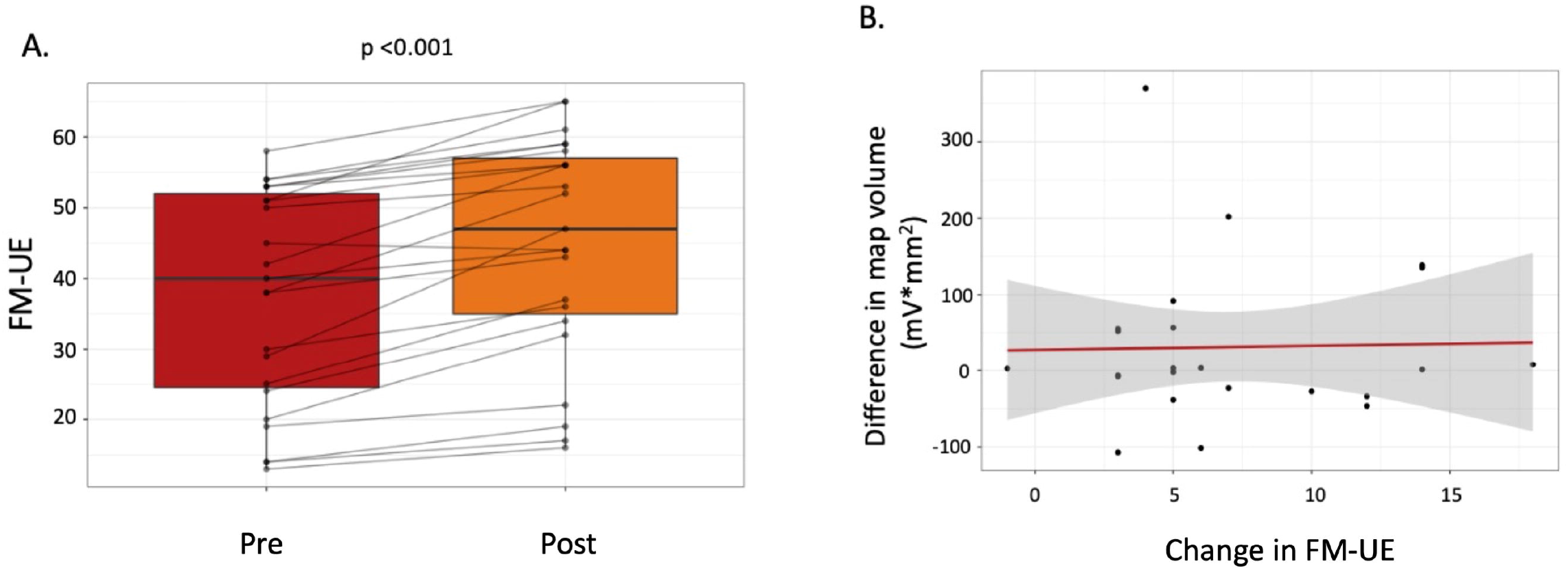
(A) The primary outcome of motor impairment (FM-UE) improved significantly with intervention, but (B) there was no correlation between the clinical improvement and changes in motor map volume of the affected side (Spearman’s rho = 0.06, p=0.79).

### TMS motor map changes

Motor threshold remained stable for Pre to Post. At the group level, ipsilesional motor map volume did not significantly change from Pre to Post (Table 1). However, we observed high inter-individual variability in map volume changes, with an increase in map volume in some participants whereas others showed a decrease, despite clinical improvements (Figure 2). There was also no significant difference in map volume changes between clinical responders (n=16) vs. non-responders (n=7) (22.97± 80.82 vs. 51.19±150.38 mV*mm^2^, p >0.9) or those who had cortical involvement (n=12) vs. subcortical only (n=11) (48.41±132.26 vs. 13.17±61.64 mV*mm^2^, p >0.9).

**Figure 2.**
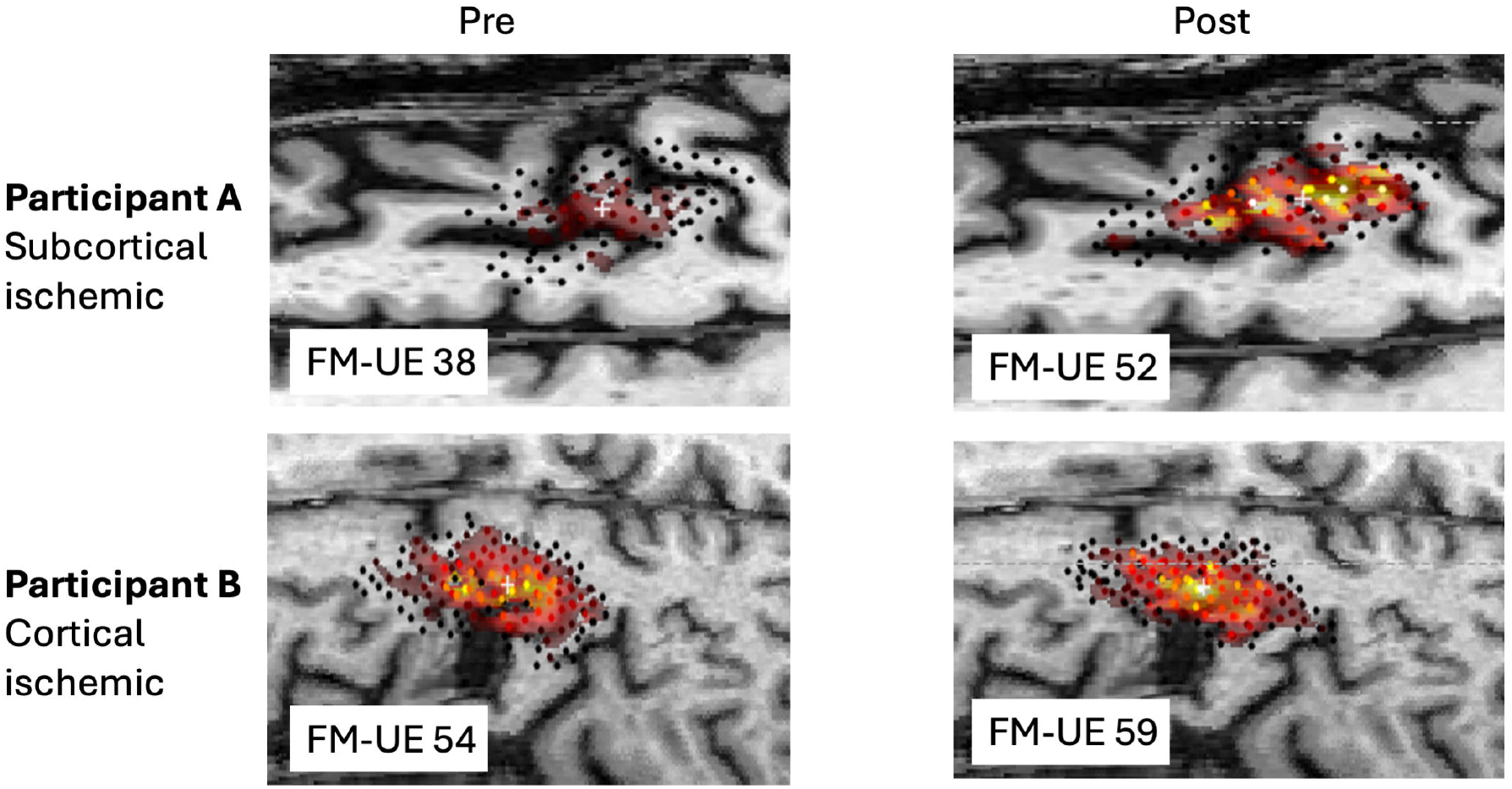
Examples of motor maps before and after intervention in the ipsilesional hemisphere for two participants.

We found no significant changes in motor map volume in the contralesional hemisphere between Pre and Post (Table 1).

### Correlation of clinical changes with TMS map changes

FM-UE changes were not significantly correlated with changes in ipsilesional map volume (Spearman’s rho = 0.06, p = 0.79, Figure 1B). We also examined the correlation of map changes with improvements on our secondary outcome measures (ARAT and WMFT) that assess functional tasks. There was no significant correlation between changes in ipsilesional map volume and ARAT (Spearman’s rho 0.36, p=0.09), or WMFT (Spearman’s rho 0.28, p=0.20).

## DISCUSSION

The significant clinical improvement across three accepted outcome measures spanning impairment and activity, was not accompanied by a corresponding increase in TMS motor map volumes. This finding held irrespective of whether the lesion affected cortex or was confined to subcortical damage. Corticospinal excitability (measured by RMT) also did not significantly change in the presence of clinical improvement.

While the motor map probably reflects motor function of the upper limb at some level, clinical improvements may not solely rely on consistent changes in the cortical motor representation as measured following stimulation at the scalp. Local intracortical network changes in synaptic weighting may have accompanied improved function, that may not be captured by overall map volume.

There are other considerations in the interpretation of our results. First, due to the timing of our assessments 7 weeks after beginning of the intervention, we may have missed map changes that occurred earlier. Second, we used one muscle (EDC) for mapping, whereas the clinical outcomes were a global measure of upper limb movement, not limited to movements directly involving the EDC muscle. It is possible that we would have observed a correlation with map changes with a more direct measure of EDC function. However, others have demonstrated that corticospinal tract integrity to distal muscles inform recovery of the entire upper limb^12^. Third, the data collection technique relied on a neurosurgical mapping method with single pulses at each stimulation location with greater spatial resolution, rather than taking an average of multiple MEP responses at points that are spaced apart. This could have influenced our findings due to the inherent variability of MEP responses^13^. Finally, here we used a TMS mapping intensity of 110% RMT, while contemporary methods recommend considerably higher (>150% RMT)^14^.

## CONCLUSIONS

Improved upper limb movement associated with intensive behavioral intervention post-stroke can occur in the presence of stable corticospinal excitability and without consistent changes in motor map representation of the finger muscles. Future studies that examine longitudinal map changes with interventions should consider timing and the effect of different mapping techniques. Furthermore, the role of M1 cortical representation may need to be examined with other structural and functional brain investigational methods in the context of broader network of recovery of upper limb motor function.

## Data Availability

All data produced in the present study are available upon reasonable request to the authors.

## ACKNOWLEDGMENTS

We would like to acknowledge Alasdair C. McLean, Samuel J. Stephen, Alex G. Chalco, and Usman M. Arshad for development of the mapping analysis software.

