## Supplemental Materials for "The human cortical motor map in post-stroke hemiparesis in the presence of treatment-related clinically important recovery"

**Supplemental Material**

Neurophysiological Outcome Measures – TMS

*Subject position and equipment*

Patients were seated in a supine position in a reclining chair, with both hands resting pronated in a pillow. Surface electrodes (Ambu Neuroline 720 self-adhesive electrodes, model number 720 01-K) were placed over the Extensor Digitorum Communis (EDC) muscles bilaterally. The signal was amplified (gain 450-550) and filtered (10-500Hz), then sampled at 1kHz using Nexstim NBT software. TMS was performed using a NBT Nexstim TMS II Stimulator with a figure-of-eight coil (230µs biphasic pulse, NBT 1.2.0 software).

*Resting Motor Threshold Procedure*

Prior to any brain stimulation, the 3D visual brain reconstruction was peeled to 22.5mm depth. Using real-time neuro-navigation, the coil was positioned over the hand knob of the contralesional hemisphere primary motor cortex (M1) and repeated for the ipsilesional hemisphere (or where the hand-knob did not exist, the perilesional cortex). Stimulation intensity was adjusted to an E-field value of 80-100 V/m. The EDC optimal site estimation was performed by delivering several supra-threshold pulses along the central sulcus on the hand knob until responses diminished. Subsequently, several pulses were delivered anterior and posterior to the sulcus adjacent to the stimulation sites that elicited the highest amplitude motor-evoked potential (MEP) in the EDC. Throughout the procedure, pulses were delivered to be perpendicular to the sulcus and in the anterior direction. The optimal site was determined as the site with the highest amplitude response from the EDC. Motor threshold (MT) was measured from the optimal site via the Nexstim NBT software Motor Threshold Determination tool. Throughout brain stimulation procedures, it was ensured that the subject’s EDC remained at rest. During MT determination, any active MEP (defined as EMG activity >25µV within 100ms prior to stimulus onset) or artifact was rejected by the TMS operator using the right foot pedal immediately following the stimulus.

*Mapping procedure*

After determining MT for the EDC, the stimulator was set to 110% MT, and the mapping procedure was initiated. Using the NBT software, a “target” was placed on the location of the rough hotspot estimation. This target was set as “active”, which overlaid a targeting grid (3mm x 3mm squares) over the surface of the cortex, providing an organizational framework for mapping. The mapping was initiated by delivering the first pulses along the central sulcus on the hand knob, with pulses being delivered perpendicular to the sulcus, the orientation of the front of the coil being at roughly a 45-degree angle to the midline, and the stimulation in an anterior direction. The operator delivered pulses along the central sulcus in both the inferior and superior directions until responses diminished. At least one pulse was delivered in each square of the grid that was mapped, and an effort was made to elicit at least two valid resting MEP’s per square of the grid. Subsequently, the operator continued to map in the anterior and posterior directions, delivering pulses at adjacent squares in all directions until responses diminished, and filling in any squares in between where stimuli had not been delivered as previously described. The result was a motor map (all “squares” of the mapping grid filled in) of the EDC representation in M1, bordered by stimuli that had elicited no responses.

After completing the motor map of the contralesional hemisphere, new electrodes were attached in the same fashion to the paretic APB and EDC, and the mapping procedure was repeated on the ipsilesional hemisphere. Special care was taken when mapping the ipsilesional hemisphere by delivering at least 3 pulses to a stimulation site (individual square of mapping grid) in the case that it was adjacent to a lesion. In the case that there was no valid resting MEP elicited in the ipsilesional hemisphere at the stimulus intensity eliciting the desired e-field value of 80-100 V/m, stimulus intensity was increased up to 100% maximum stimulator output (MSO) in order to attempt to elicit a resting response. In the case that no appreciable responses were elicited at rest, several stimuli were delivered around the area of the hand knob in order to demonstrate the absence of an elicited response from the cortex.
